# The human vestibular cortex: functional anatomy, connectivity and the effect of vestibular disease

**DOI:** 10.1101/2021.07.22.21260061

**Authors:** Richard T. Ibitoye, Emma-Jane Mallas, Niall J. Bourke, Diego Kaski, Adolfo M. Bronstein, David J. Sharp

**Affiliations:** Computational, Cognitive and Clinical Neuroimaging Laboratory, Department of Brain Sciences, Imperial College London, London, UK; Neuro-otology Unit, Department of Brain Sciences, Imperial College London, London, UK; UK Dementia Research Institute, Care Research & Technology Centre, Imperial College London, London, UK; Department of Clinical and Motor Neurosciences, Centre for Vestibular and Behavioural Neurosciences, University College London, London, UK; Centre for Injury Studies, Imperial College London, London, UK

## Abstract

Area OP2 in the posterior peri-sylvian cortex has been proposed to be the core human vestibular cortex. We defined the functional anatomy of OP2 using spatially constrained independent component analysis of functional MRI data from the Human Connectome Project. Ten distinct subregions were identified. Most subregions showed significant connectivity to other areas with vestibular function: the parietal opercula, the primary somatosensory cortex, the supracalcarine cortex, the left inferior parietal lobule and the anterior cingulate cortex. OP2 responses to vestibular and visual-motion were analysed in 17 controls and 17 right-sided unilateral vestibular lesion patients (vestibular neuritis) who had previously undergone caloric and optokinetic stimulation during functional MRI. In controls, a posterior part of right OP2 showed: (a) direction-selective responses to visual motion; and (b) activation during caloric stimulation that correlated positively with perceived self-motion, and negatively with visual dependence. Patients showed abnormal OP2 activity, with an absence of visual or caloric activation of the healthy ear and no correlations with dizziness or visual dependence – despite normal brainstem responses to caloric stimulation (slow-phase nystagmus velocity). A lateral part of right OP2 showed activity that correlated with chronic dizziness (situational vertigo) in patients. Our results define the functional anatomy of OP2 in health and disease. A posterior subregion of right OP2 shows strong functional connectivity to other vestibular regions and a visuo-vestibular profile that becomes profoundly disrupted after vestibular disease. In vestibular patients, a lateral subregion of right OP2 shows responses linked to the challenging long-term symptoms which define poorer clinical outcomes.

**Significance statement:** The human cortical vestibular network is critical to higher vestibular functions such as the perception of self-motion, judgements about verticality (‘which way is up’), and adaptation following peripheral vestibular disease (e.g. vestibular neuritis). The functional organisation of this network has remained poorly understood. In this study, we define the functional anatomy of area OP2 - a core region within the human cortical vestibular network. We identify subregions of OP2 with strong connectivity to other cortical vestibular areas. We show specific subregions of right OP2 process vestibular and visual motion information in health and that such processing is disrupted following peripheral vestibular disease. Abnormal signal processing within OP2 may underpin chronic dizziness following peripheral vestibular disease.

## Introduction

The cortical vestibular system is fundamental to self-motion perception and balance (Cullen, 2019). Area OP2 in the posterior peri-sylvian region is a core vestibular area in humans (Eickhoff et al., 2006b, 2006c; Lopez et al., 2012; zu Eulenburg et al., 2012), but its functional organisation remains unclear.

Area OP2 is located in the posterior parietal operculum and extends into the retroinsular (posterior insular) cortex (Fig. 1A) (Eickhoff et al., 2006c). Meta-analyses have shown consistent activation in OP2 following vestibular stimulation (Lopez et al., 2012; zu Eulenburg et al., 2012). OP2 is homologous to the non-human primate PIVC - a ‘core’ cortical vestibular region (Eickhoff et al., 2006c) receiving vestibular, visual and somatosensory inputs (Guldin and Grüsser, 1998). Neural activity in non-human primate PIVC encodes head motion (Grüsser et al., 1990a, 1990b; Chen et al., 2010) and is necessary for accurate perception of self-motion (Chen et al., 2016).

**Figure 1.**
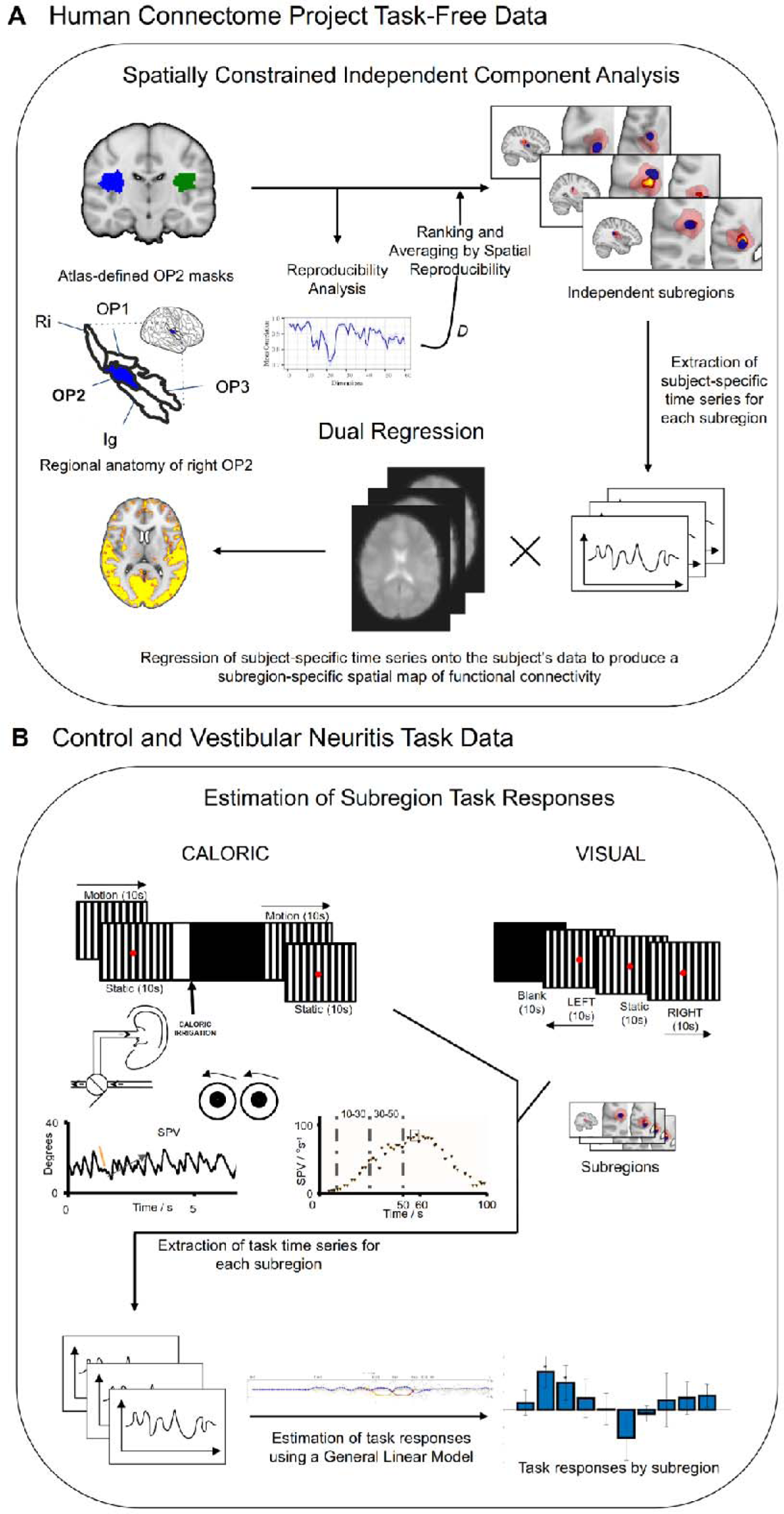
Overview of methods. (A) - Spatially-constrained independent component analysis of area OP2 within Human Connectome Project task-free data produced spatial maps of subregions in each hemisphere. Between-subject reproducibility was maximised and reproducibility was further enhanced by Ranking and Averaging by Spatial Reproducibility. The whole brain connectivity of each subregion was then estimated by dual regression. The regional anatomy of right OP2 (in blue) is shown on an inflated brain relative to areas OP1, OP3, the retroinsular cortex (Ri) and the insular granular cortex (Ig); the images are adapted with permission from Indovina et al. (Indovina et al., 2020) (B) CALORIC and VISUAL tasks were performed. The trace on the left is an example of right-beating nystagmus during cool caloric irrigation of the left ear; the orange line depicts slow phase velocity (SPV) which is defined as the gradient of the slow phase of nystagmus; the adjacent trace is a typical profile of the evolution of slow phase velocity over time following caloric irrigation at time zero; the time periods (10 – 30 seconds, and 30 – 50 seconds) which define the caloric contrast are illustrated.

Areas neighbouring OP2 respond to visual as well as vestibular stimuli (Brandt et al., 1998; Sunaert et al., 1999; Frank et al., 2014, 2016b). The retroinsular cortex is proposed to integrate visual and vestibular motion information, whereas more anterior areas in OP2 process vestibular information (Frank and Greenlee, 2018). The processing of visual and vestibular information is affected by unilateral vestibular failure, e.g. vestibular neuritis - a major cause of acute vertigo. Despite central adaptation, many patients are left with long-term dizziness (Imate and Sekitani, 1993; Cousins et al., 2014) and the failure to recover fully is accompanied by over-reliance on visual information for perceptual and balance judgements (visual dependence) (Kanayama et al., 1995; Bronstein and Dieterich, 2019). The neural basis of this change is unknown but based on data showing an association between grey matter volume in OP2 and symptom load (Helmchen et al., 2009), and altered functional connectivity in OP2 neighbouring regions following vestibular neuritis (Helmchen et al., 2014), we propose neural activity within OP2 is relevant to multisensory processing.

An understanding of the vestibular function of OP2 could be informed by studying its functional connectivity. Structurally connected regions often show correlated brain activity, which reflects shared functions (Smith et al., 2009). Within the brain, subregions with different functions can be identified by their distinct patterns of functional connectivity using spatially constrained independent component analysis (ICA) (Leech et al., 2011, 2012; Moher Alsady et al., 2016). This has been an informative approach for understanding the functions of a range of brain regions (Leech et al., 2012; Moher Alsady et al., 2016; De Simoni et al., 2018), but the method has never before been applied to the vestibular cortex.

Thus, the main aim of this study is to define the functional anatomy of OP2 in a data-driven way using spatially constrained ICA applied to Human Connectome Project (HCP) data (Glasser et al., 2013). In addition, using data collected during a previous study on visual-vestibular activation (Roberts et al., 2018) we investigate OP2 responses in controls and patients with chronic vestibular neuritis to see: (i) whether OP2 activation during caloric irrigation and visual motion stimulation is homogeneous or restricted to specific functional subregions; (ii) if the latter is true, calorically activated OP2 subregion(s) should show connectivity to other known vestibular areas; (iii) ascertain whether OP2 underpins perceptual functions so that activation during caloric stimulation correlates with perceived self-motion (dizziness), visual dependence, and clinical outcome (chronic dizziness) in vestibular neuritis; and (iv) OP2 responses to caloric irrigation are altered in chronic vestibular neuritis as a corollary of central adaptation.

## Materials and Methods

### Participants

The Human Connectome Project is an openly accessible dataset of high-quality neuroimaging data (Glasser et al., 2013). MRI data from 100 healthy unrelated participants (54 females, aged 22-36 years) within the Human Connectome Project (HCP) were used.

Seventeen right-handed patients with chronic (> 6 months since symptom onset) right-sided vestibular neuritis participated in the original study (mean age 58.8 +/- 17.3 [SD] years, 9 female) (Roberts et al., 2018). They presented acutely with vertigo. Examination revealed left-beating horizontal jerk nystagmus and catch-up saccades following head impulse tests to the right (Halmagyi and Curthoys, 1988). Caloric testing at diagnosis confirmed paresis of the right horizontal canal (mean 53.1 %, SD 34.1 %). Patients had no other vestibular symptoms or pre-existing vestibular or neurological disorders. Audiometry was normal for age. The burden of dizziness handicap, the frequency of dizziness symptoms, and the extent of visually induced dizziness were captured using questionnaires: the Dizziness Handicap Inventory (DHI), Vertigo Symptom Scale (VSS) and Situational Vertigo Questionnaires (SVQ) respectively (Jacob et al., 1989; Jacobson and Newman, 1990; Yardley et al., 1992). In the original study (Roberts et al., 2018) 17 right-handed healthy age-matched controls were recruited (mean age 58 +/- 14 [SD] years, 10 female). They had no history of vestibular or neurological disorders. Caloric vestibular test results were normal. Written informed consent was obtained from all participants. All procedures were performed in accordance with the ethical standards of the Bromley and the Fulham local research ethics committee.

### Psychophysical measures

Patients with vestibular neuritis and controls provided dizziness ratings (Likert scale 0 to 10, higher was more dizzy). These ratings reflected the intensity of perceived self-motion immediately after each caloric task functional MRI session. All participants had a screening caloric test prior to the functional MRI sessions; their subsequent dizziness ratings were in comparison to the screening experience which was rated ‘5’.

Visual dependence was measured outside the scanner by the extent to which the subjective visual vertical was biased by background visual motion (Fig. 7C(ii)) (Cousins et al., 2014; Roberts et al., 2017). Here, an estimate of the perceived vertical – the subjective visual vertical, was first measured. This was then repeated with background visual motion (Fig. 7C(ii)). Participants looked at a computer screen through a 30 cm deep viewing cone while standing. A 6 cm white rod was presented on a black background. Around a central 6 cm circle was a field of 220 randomly distributed off-white dots each subtending 1.5 degrees of visual field. The rod was rotated to a random angle then participants were asked to re-align it to their subjective visual vertical using keyboard controls. The procedure was then repeated during clockwise and counter-clockwise rotation of the disc. Each condition was repeated 6 times, then results were averaged. Visual dependence was defined as the absolute bias of verticality estimates in the motion conditions minus the subjective visual vertical with a static background.

### MRI acquisition and preprocessing

HCP MRI data had been collected on a 3T Siemens Connectome Skyra scanner and included a T1-weighted MPRAGE image and a resting-state functional MRI sequence for each participant (Glasser et al., 2013). HCP MRI data had been optimally pre-processed by the HCP consortium with FMRIB Software Library (FSL) and Freesurfer (version 52) tools. Task-free functional (echo-planar) MRI data were registered to Montreal Neurological Institute (MNI) standard space in a two-step process; a linear transform of the participant’s echo-planar image to their structural (T1-weighted) image was combined with a non-linear transform of the participant’s structural image to the standard (MNI) structural image (Jenkinson et al., 2002). A high-pass filter (2000 seconds) was then applied. Artefacts were removed using FMRIB’s ICA-based X-noisifier (FIX) (Salimi-Khorshidi et al., 2014). FIX classifies components and regresses out noise and motion-related signals.

Anatomical and functional MRI data from patients with vestibular neuritis and controls were acquired on a Siemens Verio 3T scanner using standard procedures (Roberts et al., 2017). Echo-planar functional T2*-weighted imaging data were acquired in 44 axial planes using a gradient echo sequence (interleaved order, TR 2500 ms, TE 30ms, flip angle 80 degrees, voxel dimension 3 x 3 x 3 mm, acquisition matrix 64 x 64). Foam pads were used to minimise head movement. T1-weighted images were also acquired (TR 2300 ms, TE 3 ms, TI 900 ms, flip angle 9 degrees, bandwidth 238Hz/pixel, voxel dimensions 1 x 1 x 1 mm, matrix size 256 x 192, FOV 240 x 256 mm, number of excitations = 1).

MRI data from patients with vestibular neuritis and controls were preprocessed using FSL tools. Structural images were brain-extracted using the Brain Extraction Tool (Smith, 2002). Echo-planar images were registered to Montreal Neurological Institute (MNI) standard space in a two-step process; a linear transform of the participant’s echo-planar image to their structural (T1-weighted) image was combined with a non-linear transform of the participant’s structural image to the standard (MNI) structural image (Jenkinson et al., 2002). Images were spatially smoothed (6 mm full-width at half-maximum Gaussian kernel) then high-pass filtered (100 seconds). Motion was addressed by motion-correction, and denoising using ICA. Estimates of motion within echo-planar images were determined to 6 degrees of freedom using MCFLIRT within FSL (Jenkinson et al., 2002). These estimates were then applied as nuisance regressors to derive motion-corrected images. Preprocessed MRI data were also decomposed using ICA, and motion components were removed using an ICA-based tool for the Automatic Removal of Motion Artifacts (ICA-AROMA) (Pruim et al., 2015). Data were pre-whitened prior to analysis within general linear models (Woolrich et al., 2001).

### Caloric and visual tasks

Patients with vestibular neuritis and controls underwent: (i) a caloric protocol (four conditions of caloric irrigation flanked by visual stimulus blocks); and (ii) a visual localiser task (Roberts et al., 2018). The caloric protocol consisted of cool or warm caloric irrigation in the left ear flanked by 60 second visual motion stimulus blocks with leftward or rightward motion (Fig. 1B), giving a total of four conditions. The visual localiser task was performed during a separate functional MRI session (Fig. 1B) and consisted of a 40 second block of stimuli including left or right moving gratings, repeated six times, totalling 240 seconds. A description of the tasks is presented in Fig. 1B.

### Caloric protocol

Participants were instructed to keep their eyes open throughout. The direction of visual motion stimuli within each block were either left or right. The first visual motion stimulus block comprised 10 seconds of a horizontally-moving vertical grating, followed by 10 seconds of no motion (‘static’), repeated three times, during which participants were instructed to look at a central fixation dot subtending 0.5 degrees. The grating was composed of black and white vertical bars. Each vertical stripe subtended an angle of 1.9 degrees, with the whole screen subtending a total angle of 15 degrees. During periods of motion the visual grating moved leftward or rightward at 8 degrees per second. After the visual motion stimulus block, there were 5 seconds of a written instruction: ‘Get Ready’, then a 2.5 second written instruction to ‘turn on the tap’, following which participants initiated caloric irrigation by turning a tap by hand (Fig. 1B) (Roberts et al., 2017, 2018). The irrigation lasted 50 seconds. During irrigation, the screen was black. Two hundred and fifty millilitres of cool (30 °C) or warm (44 °C) water flowed into the left external auditory canal. Irrigation was followed by a second visual motion stimulus block of the same composition as the first. Eye movements were recorded throughout by an infrared MRI-compatible eye tracker (Ober consulting, Poland).

The direction of slow phase eye movements induced by caloric irrigation depends on caloric temperature such that a warm stimulus in the left ear causes the eyes to drift rightwards. The direction of the final visual block’s motion stimuli combined with warm or cool caloric stimuli to produce four conditions which were either congruent or incongruent regarding the consistency of the directions of slow phase eye movements and visual motion (Congruent: cool irrigation + left visual motion [CL], and warm irrigation + right visual motion [WR]; Incongruent: cool irrigation + right visual motion [CR], and warm irrigation + left visual motion [WL]) (Roberts et al., 2017, 2018).

### Visual localiser task

The visual localiser task was performed during a separate functional MRI session (Fig. 1B). A 40 second block of stimuli was repeated six times, totalling 240 seconds. Each stimulus block consisted of 10 seconds of a black screen followed by 10 seconds of a leftward moving black and white vertical grating, 10 seconds a static grating, then finally 10 seconds of a rightward moving grating (Roberts et al., 2017, 2018). The visual grating had the same characteristics as that used in the caloric protocol visual motion stimulus blocks.

### MRI analysis

#### Spatially constrained independent component analysis

Spatially constrained ICA was applied to OP2 in the HCP task-free functional MRI dataset, using the masked ICA (mICA) toolbox (Moher Alsady et al., 2016). Right and left OP2 masks were created using the Jülich probabilistic atlas within the FMRIB Software Library (FSL) (Eickhoff et al., 2006a). The masks were not thresholded to ensure immediately adjacent parietal opercular and retroinsular areas were included. Masked participant data was spatially smoothed using a 5 mm kernel prior to ICA.

#### Split-half sampling

To select an optimum dimensionality for the ICA decomposition in a data-driven way, we undertook split-half sampling as implemented in the mICA (Moher Alsady et al., 2016). This method maximises between-subject reproducibility. The analysis of between-subject reproducibility involved partitioning the HCP participants into two halves by random sampling, repeated 50 times. Group ICA was then undertaken for each of the two halves over 1 to 60 dimensions, for each sampling repetition. Independent components with similar functional connectivity were matched by the Pearson correlation coefficient of their time series between the two split-halves. The mean correlation of matched time series pairs across repetitions was the measure of reproducibility. As recommended, we selected the dimensionality with the highest global reproducibility (Moher Alsady et al., 2016). The ideal number of dimensions for ICA was first established in right OP2. Left OP2 was then decomposed into the same number of subregions. The motivation for starting with right OP2 is the meta-analysis evidence which shows right OP2 is core to the cortical vestibular response (zu Eulenburg et al., 2012).

#### Ranking and Averaging Independent Component Analysis by Reproducibility

ICA results are unstable across repeat decompositions (realisations), limiting reproducibility (Himberg et al., 2004). We therefore applied an established method where the results of multiple realisations were ranked and averaged by their spatial reproducibility (Ranking and Averaging Independent Component Analysis by Reproducibility, RAICAR (Yang et al., 2008), source code available at https://github.com/yangzhi-psy/RAICAR). ICA was repeated 30 times for the dimensionality which maximised between-subject reproducibility. Components from each realisation were then iteratively matched with the most similar components from other realisations, by maximising spatial correlation. The results of this process were distinct, aligned component sets. Each set contained members from different realisations. For each aligned component set, a cross-realisation spatial cross-correlation matrix was then produced. A default cut-off spatial correlation of 0.5 was used in this study to identify reproducible components (Yang et al., 2008). Finally, for each aligned component set, a selective average of the spatial maps and time series of the members across realisations was produced. Only realisations which had at least one spatial correlation coefficient higher than the cut-off threshold were included in this selective average.

### Dual regression

We investigated the whole brain connectivity of OP2 subregions as derived from HCP data. To do this, we applied dual regression within FSL (Fig. 1A). Subregion spatial maps were applied to HCP task-free data to derive participant-specific time series which were then regressed onto participant functional MRI data to produce participant-specific spatial maps of functional connectivity for each subregion (Nickerson et al., 2017). Group-level maps for each subregion were then derived by non-parametric permutation testing using *randomise* (within FSL, n =10,000, p < .05) (Braga et al., 2013). We tested for the inclusion of known regions of interest from zu Eulenburg et al.’s meta-analysis of vestibular stimulation studies, as evidence of vestibular connectivity (zu Eulenburg et al., 2012).

### Caloric and visual task responses

We investigated the response of OP2 subregions during caloric stimulation using an established approach (Leech et al., 2012; Braga et al., 2013). Here, the time series unique to each subregion was determined. To do this, each subregion identified by spatially constrained ICA of the HCP data was spatially regressed onto caloric task session data with the remaining subregions as nuisance covariates – the first stage of dual regression within FSL (Jenkinson et al., 2002; Nickerson et al., 2017). The resulting subregion-specific time series for each session then served as inputs to general linear models to estimate responses using contrasts (Leech et al., 2012).

Mirroring the original methodology applied to this data, vectors representing the onsets of visual motion, visual static and caloric irrigation were convolved with a double-gamma haemodynamic response function and its temporal derivative (Roberts et al., 2017). Peak vestibular activation was modelled using a contrast of vectors representing the last 20 seconds of caloric irrigation and the previous 20 seconds (Fig. 1B). This controlled for the effects of auditory and somatosensory activation while avoiding contamination from movement in the first seconds after caloric onset (note: participants self-triggered the caloric irrigation by turning a tap). In the current study, wherein the aim was to estimate the cortical response to vestibular stimulation, the contrast of interest was the caloric response (which precedes and is independent of the second visual stimulus block, Fig. 1B). Consequently, as the post-caloric visual stimulus was not of interest, information from the four conditions were combined to produce a summary caloric response for each participant. For each participant, results across their four sessions were transferred to higher level models by a summary statistic approach (Beckmann et al., 2003), modelling subregion responses at the participant level, and at the group-level adjusted for linear effects of age and sex.

To determine the response of OP2 during caloric irrigation, we undertook a whole brain analysis of the control caloric task dataset within FSL, here using a higher-level mixed effects (FLAME 1 + 2) model to estimate the group response (Woolrich et al., 2001). Clusters were inferred from the resultant z-statistic images, applying a threshold of z > 3.1 (p < .001) and a corrected cluster significance threshold of p < .05. This activation was compared to a meta-analysis-derived mask of voxels, wherein at least one study showed significant activation to vestibular stimulation (Fig. 2) (Lopez et al., 2012). Meta-analysis mask data were provided courtesy of Dr. Christophe Lopez, Aix-Marseille Université (Marseille, France).

**Figure 2.**
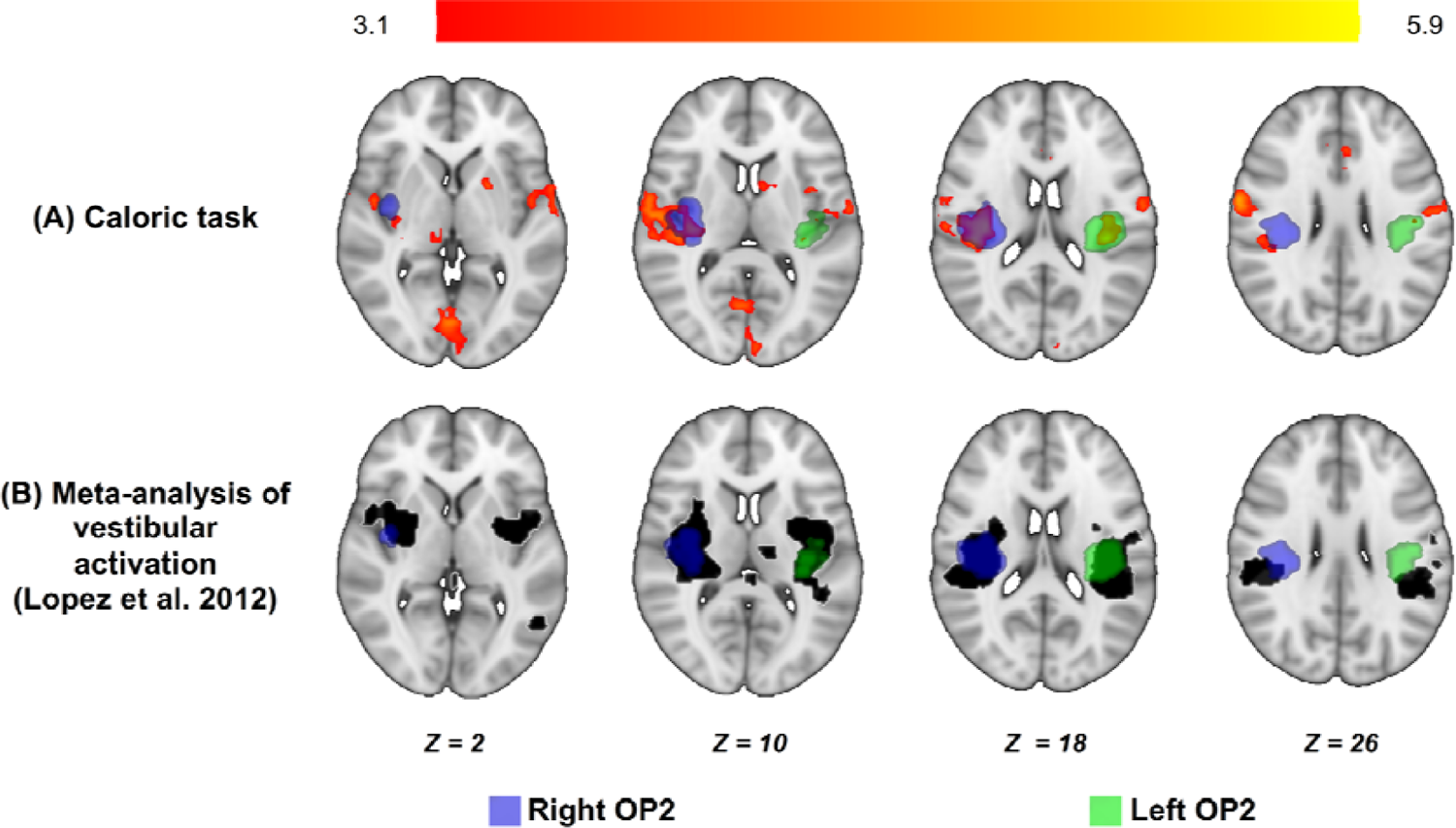
Bilateral OP2 activation during caloric irrigation. (A) Areas of significant activation during caloric irrigation in healthy controls (red-yellow). Activation is seen in right and left OP2. Masks of right (blue) and left OP2 (green) are shown, defined using the Jülich atlas. (B). Vestibular activation produced in OP2 by acoustic, caloric or galvanic stimuli (black, courtesy of Dr. Christophe Lopez, Aix-Marseille Université (Marseille, France) (Lopez et al., 2012)).

A role in integrating visual and vestibular information implies OP2 subregions may encode the direction of visual motion stimuli. Subregion activity during visual motion stimuli might therefore vary with motion direction. To test this, we analysed the visual task dataset, defining contrasts of visual motion (rightwards minus static, leftwards minus static) and motion direction (right minus left). Here, as in the analysis of caloric task data, OP2 subregions derived from HCP data were spatially regressed onto participant data to produce subregion-specific time series. The time series were then analysed within a general linear model to estimate responses linked to visual motion and motion direction.

Corrections for multiple comparisons were undertaken by controlling the False Discovery Rate (FDR) across the subregions within each hemisphere, with a significance threshold of p < .05 (Genovese et al., 2002).

### Caloric responses and vestibular function

We investigated whether self-motion perception and/or visual dependence correlated with OP2 subregion responses during caloric irrigation. As dizziness ratings were obtained after each session, but sessions differed by stimulus condition (cool or warm caloric irrigation combined with left or right visual motion during the final visual block), the relationship between the summary caloric responses and dizziness ratings was estimated using linear mixed-effects regression (using *fitlme* within MATLAB®). To visualise results, *adjusted dizziness* was determined. Adjusted dizziness captured the relationship between post-caloric dizziness ratings and the caloric response while averaging out the contributions of other predictors in the linear model (MATLAB, 2021).

To understand the correspondence between self-motion perception (post-caloric dizziness) and nystagmus responses, we investigated the relationship between these variables. The peak nystagmus slow phase velocity was measured during each caloric task session from eye movement traces. The relationship between peak nystagmus ‘slow phase velocity’ and ‘dizziness’ was investigated using linear mixed-effects regression. ‘Dizziness’ was the dependent variable, with ‘slow phase velocity’ and ‘condition’ as fixed effects, and ‘participant’ as a random effect (in a random intercept model).

In patients with vestibular neuritis, we tested for correlation between questionnaire scores of the burden of chronic symptoms and the summary caloric response of OP2 subregions, to identify subregions which may be relevant to clinical recovery following vestibular neuritis.

### Data availability

Raw data which support these results were collected at Imperial College London. These data and derived data supporting the findings in this study are available from the corresponding authors upon reasonable request.

## Results

### Bilateral OP2 activation during vestibular stimulation

Caloric stimulation produced activation of OP2 in both hemispheres in controls (Fig. 2). Right OP2 showed more extensive activation than left OP2 (right OP2 peak: x=38, y=-20, z=17, 619 voxels; left OP2 peak: x=-38, y=-26, z=18, 219 voxels). Activation associated with caloric stimulation was also seen in the right precentral gyrus, intracalcarine cortex, left putamen, right thalamus, left thalamus, right frontal pole and left lingual gyrus. OP2 has previously been shown to be activated by caloric, galvanic and acoustic stimuli. A recent meta-analysis has reported the average location of this activation, which we compared to activation produced during our caloric stimulation study (Lopez et al., 2012). A similar pattern of OP2 activation was observed with caloric stimulation and the meta-analysis of vestibular activation (Fig. 2).

### OP2 subregions show functional connectivity with other vestibular regions

We next identified distinct subregions within OP2 on the basis of their functional connectivity using spatially constrained ICA applied to task-free fMRI data from the Human Connectome Project. Ten subregions with distinct functional connectivity were identified on the right (Fig. 3A). Constraining ICA to produce ten components maximised their reproducibility (*r* = 0.976, SD 0.112). A range of connectivity patterns were observed (summarised in Table 1). Connectivity was seen to regions involved in vestibular function (zu Eulenburg et al., 2012). Most OP2 subregions were connected with a number of vestibular areas, including the insula, parietal operculum, inferior parietal cortex and premotor cortex (Table 1 and Fig. 3A). Overall, seven of ten subregions in right OP2 were connected to the majority of vestibular regions (R1, R2, R3, R4, R5, R6 and R10, Table 1). R7 largely connected to the occipital cortex. Subregions R8 and R9 had no meaningful grey matter connectivity.

**Figure 3.**
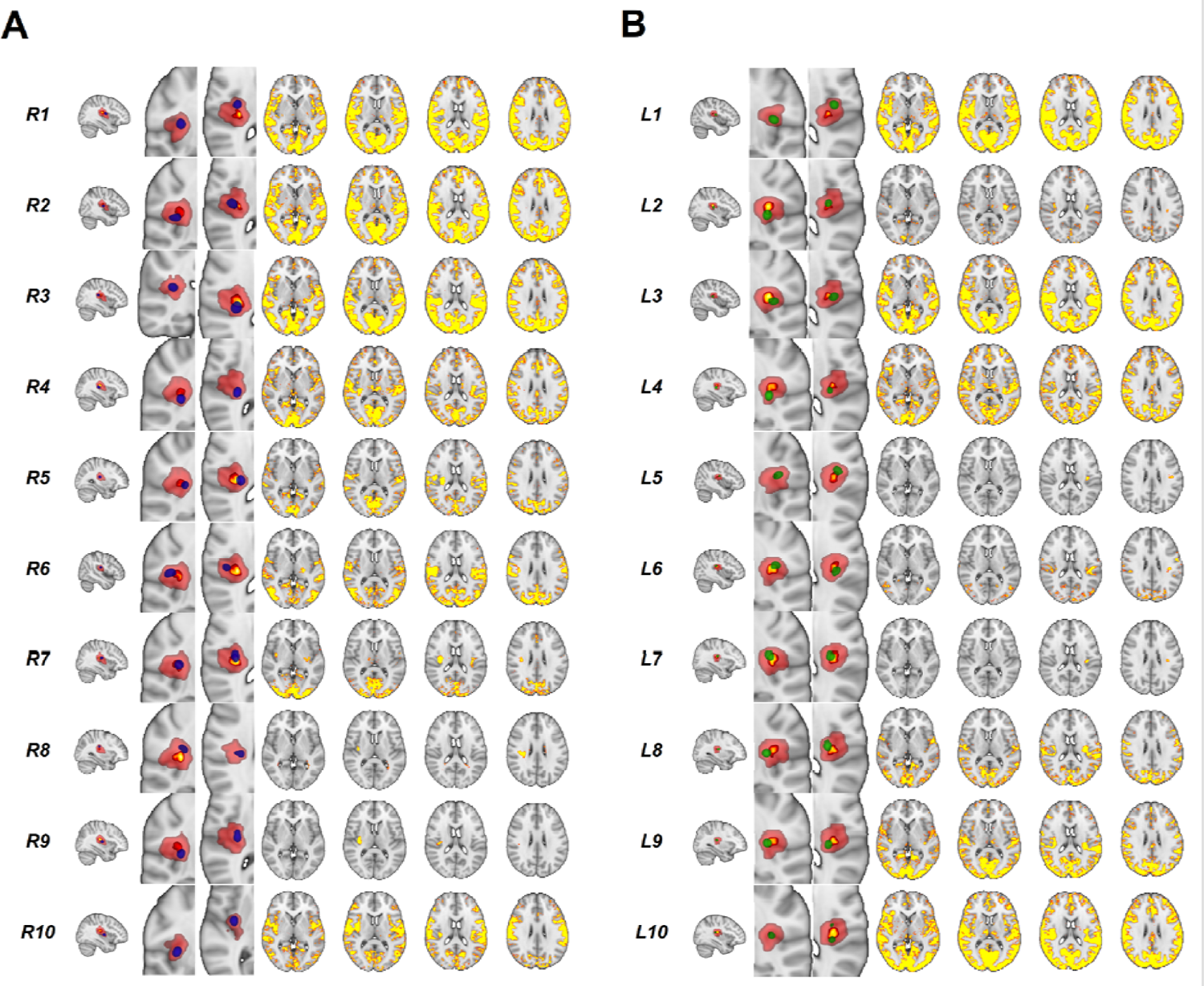
Subregions within OP2 identified from independent component analysis. (A) Right OP2 subregion images (blue) overlaid on right OP2 mask (red, from Jülich atlas). The core/central area of the OP2 mask (p > 0.5 in Jülich atlas) is shown in yellow. Brain networks with significant functional connectivity to each OP2 subregion are shown adjacent (significant voxels red-yellow, p < .05). (B) Left OP2 subregions are shown in green with adjacent functional connectivity maps. R = right. L = Left.

**Table 1.**
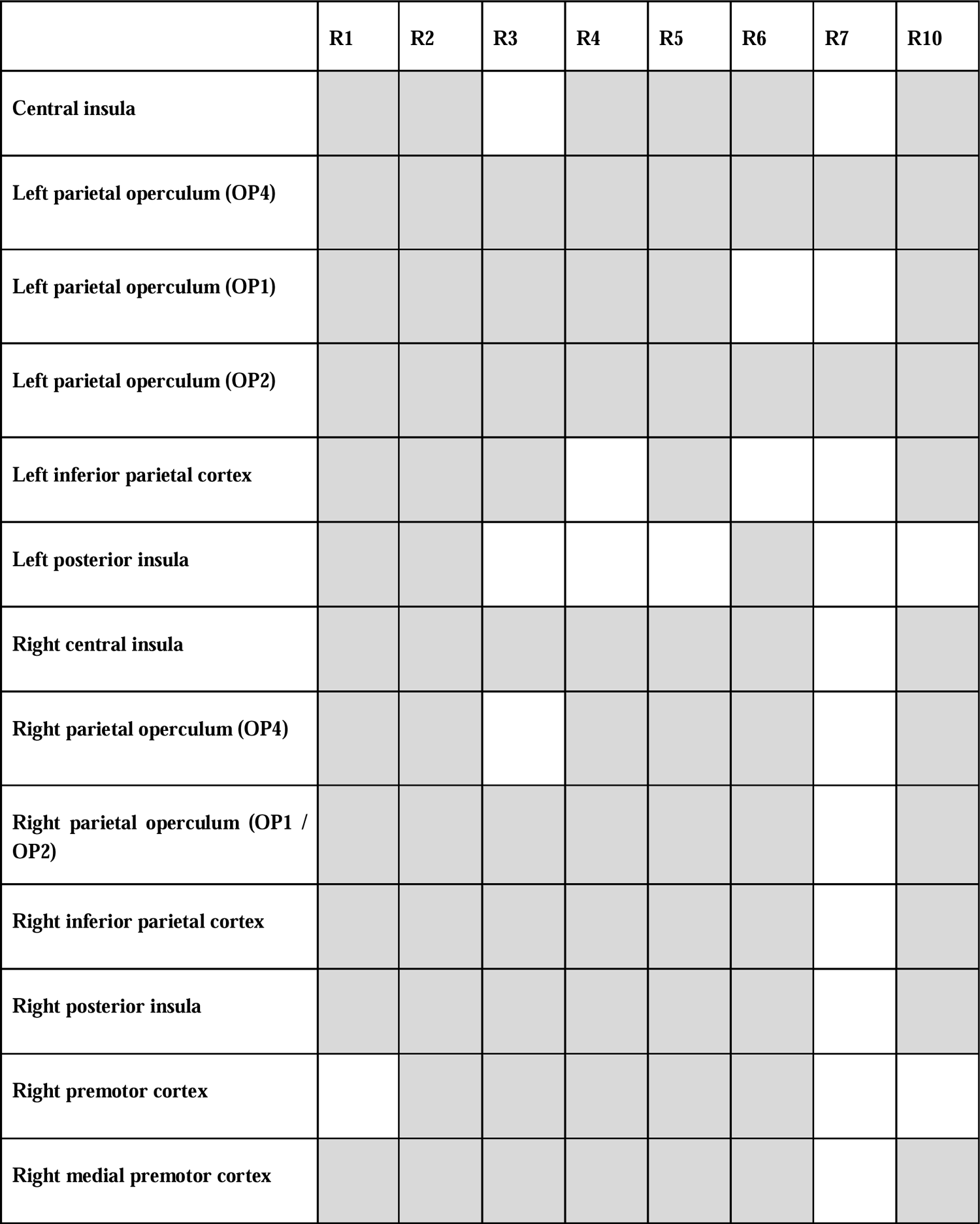
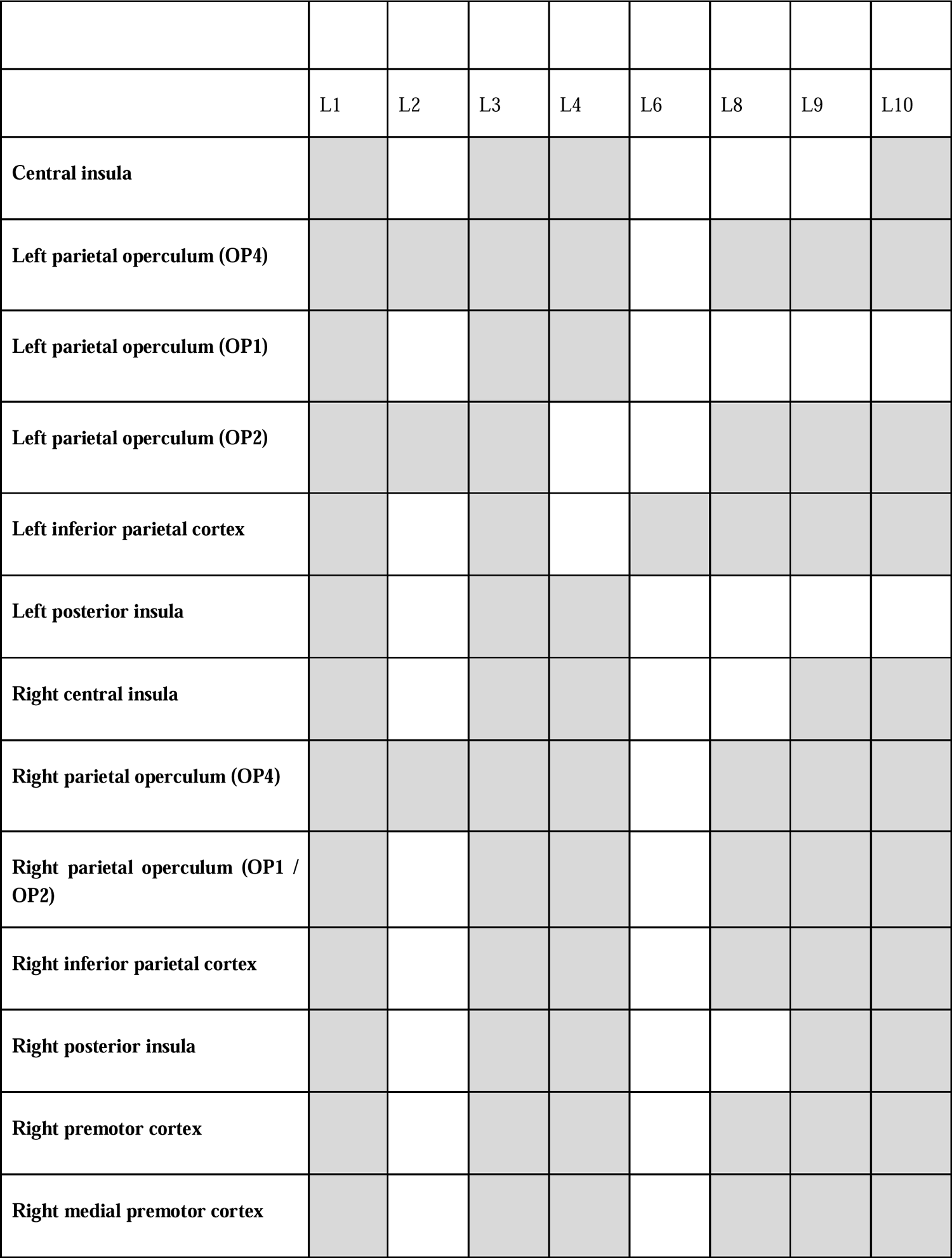
OP2 subregions are connected to known vestibular areas. . Shaded cells indicate significant functional connectivity between OP2 subregions and vestibular areas defined in zu Eulenburg et al.’s7 meta-analysis.

Ten subdivisions in left OP2 were identified in a similar way. Most subregions in left OP2 also connected to previously identified vestibular areas (Table 1). Subregions L1 and L3 connected with all vestibular regions of interest identified by meta-analysis (Fig. 3B, Table 1) (zu Eulenburg et al., 2012). Overall, six of ten subregions in left OP2 were connected to the majority of vestibular regions (L1, L3, L4, L8, L9, L10). L6 was largely connected with the occipital cortex. Subregions L5 and L7 had no meaningful grey matter connectivity (Fig. 3B).

Hierarchical clustering was used to identify spatially similar OP2 networks (Hastie et al., 2009). Three right OP2 networks had high spatial similarity to each other (R1 [anterior OP2], R2 [lateral OP2] and R3 [posterior OP2] (Dice coefficient > 0.7, Fig. 4A). Peak connectivity in this common network was with the primary somatosensory cortex, the parietal operculum (OP1 and OP2 bilaterally), supracalcarine cortex, left inferior parietal lobule and the anterior cingulate. Analysis of left OP2 connectivity showed less clear clustering of spatial similarity between whole brain connectivity networks (Fig. 4B). However, the three most similar networks on the left shared functional connectivity to a network very similar to the common network identified from right OP2 regions (Dice similarity co-efficient 0.80, Fig. 4).

**Figure 4.**
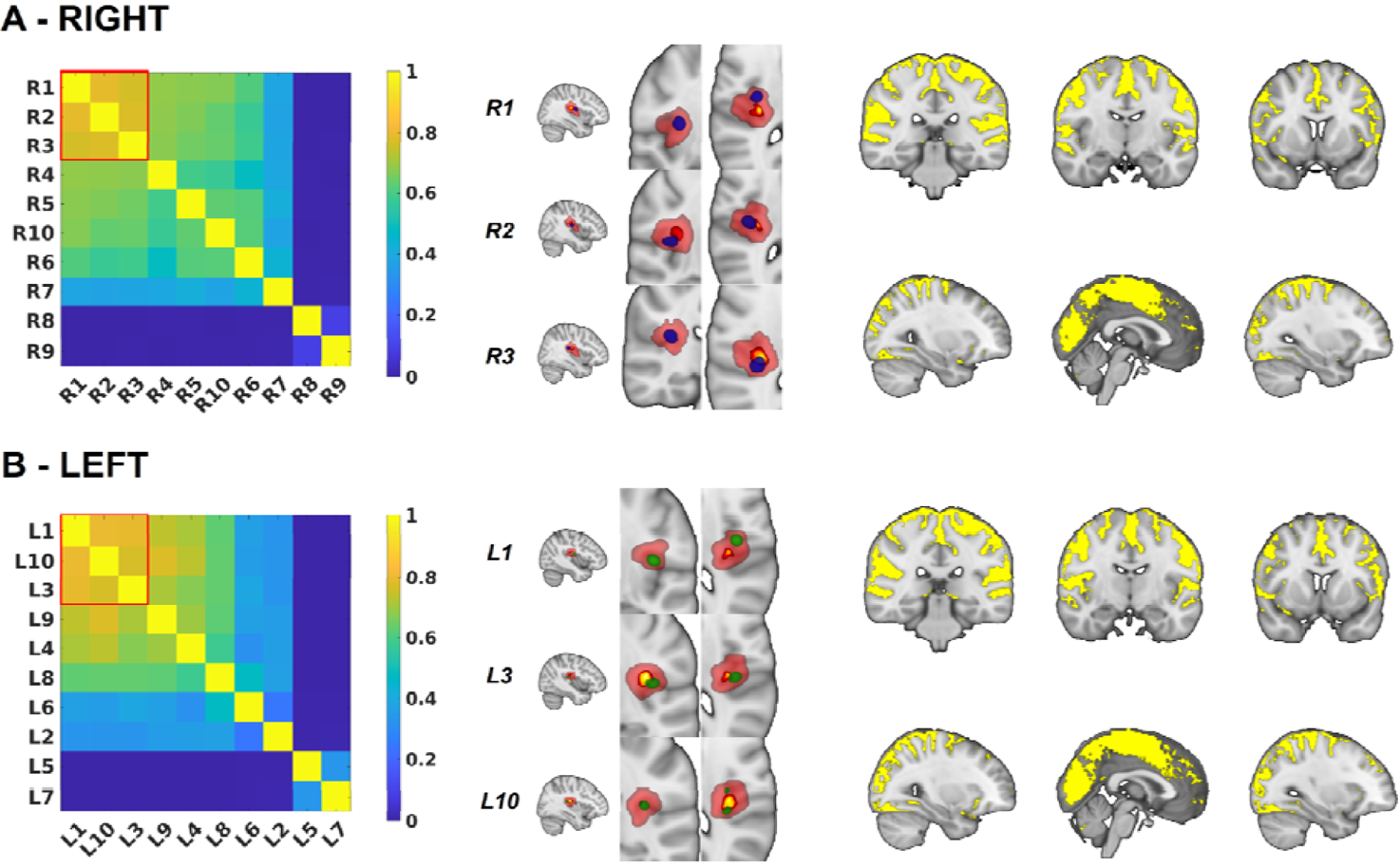
Common patterns of vestibular connectivity from OP2 subregions. (A) Spatial similarity matrix for the whole brain connectivity of right OP2 subregions as measured by the Dice coefficient. Red square highlights three subregions with similar networks (R1 [anterior OP2], R2 [lateral OP2] and R3 [posterior OP2]). These three subregions are illustrated in blue, on a background of the OP2 mask in red. The core/central area of the OP2 mask (p > 0.5 in Jülich atlas) is shown in yellow. Adjacent is a map of common functional connectivity from the three subregions (yellow, p < .05). (B) Spatial similarity matrix, OP2 subregions (in green) and common functional connectivity for left OP2.

### Area OP2 responds to caloric irrigation but not after vestibular neuritis

Next, we investigated whether specific OP2 subregions responded to caloric irrigation (of the left ear). In healthy controls, within the right hemisphere only right R3 [posterior OP2] activated significantly during left ear caloric irrigation (t(16) = 3.33, p = 0.005, FDR-corrected p < .05, Fig. 5(i)). In the left hemisphere, L3 [lateral OP2] and L10 [posterior OP2] showed significant activation (t(16) = 3.18, p = .005; L10: t(16) = 3.23, p = .006, FDR-corrected p < .05, Fig. 5(ii)). Patients with right vestibular neuritis showed abnormal vestibular responses in OP2 (Fig. 5(iii-iv)). Caloric irrigation produced no significant activation of any OP2 subregions in either hemisphere (Fig. 5(iii-iv)). R3 [posterior OP2] showed significantly lower levels of activation in vestibular neuritis patients than controls (t(32) = 2.87, p = .007). In the left hemisphere, L10 [posterior OP2] showed a reduction in activation in vestibular neuritis patients compared to controls of borderline significance (L10: t(32) = 1.98, p = .057).

**Figure 5.**
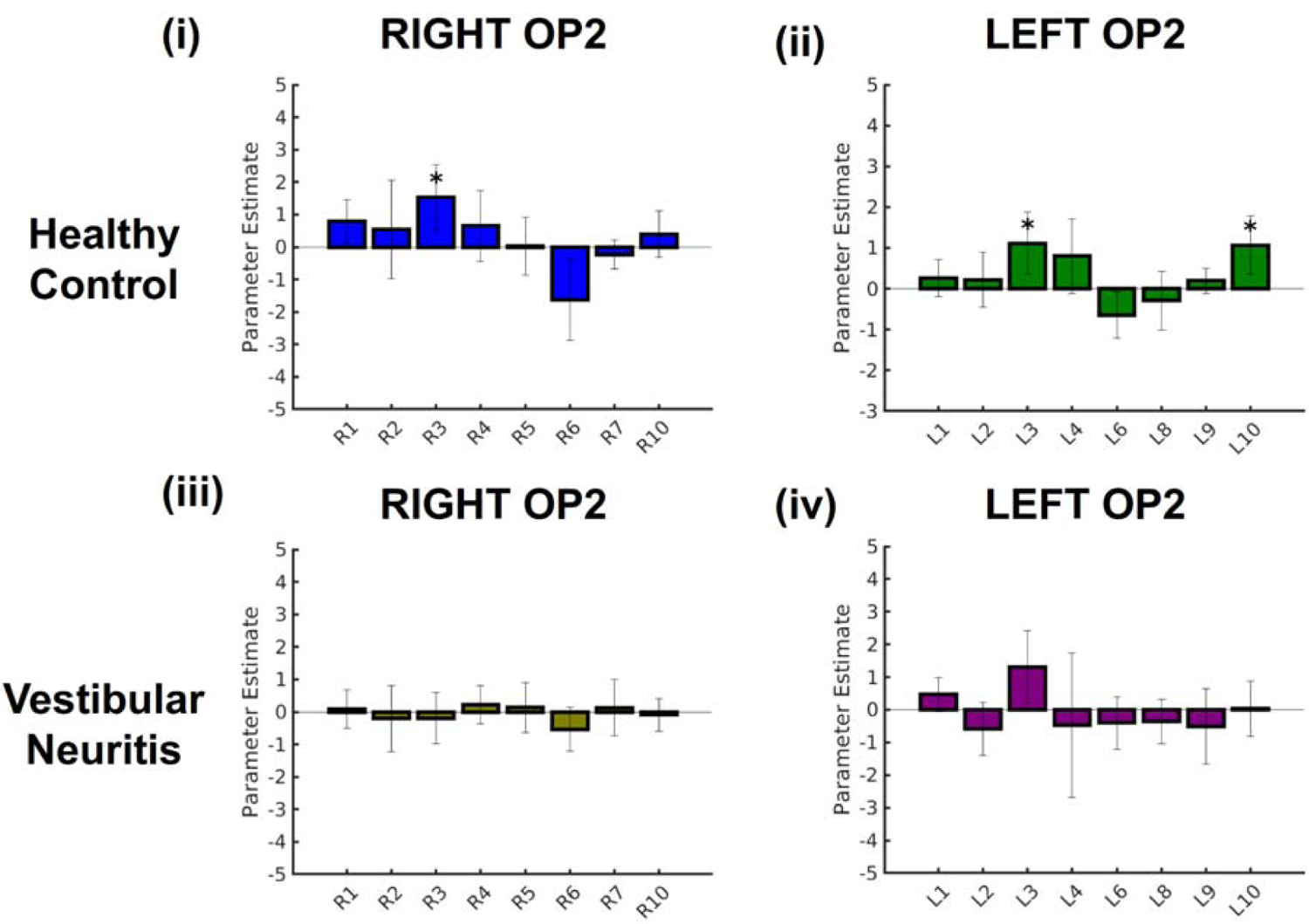
OP2 activation during caloric irrigation. Caloric responses in healthy controls (i-ii) and patients with vestibular neuritis (iii-iv) within right (i, iii) and left (ii, iv) OP2 subregions. * indicates significant activation relative to baseline, FDR-corrected.

### Right OP2 shows visual motion selectivity but not after vestibular neuritis

Right OP2 also responded selectively to the direction of visual motion in controls but not in patients with vestibular neuritis (Fig. 6). In controls, R2 [lateral OP2] activated during right visual motion and deactivated during left visual motion (right motion: t(16) = 2.41, p = .030, FDR-p > .05; left motion: t(16) = −3.28, p = .005, FDR-p < .05). Other subregions within right OP2 did not show activation significantly modulated by motion. The direct comparison of responses to left and right visual motion showed that both right R2 [lateral OP2] and R3 [posterior OP2] showed motion selectivity in controls, responding significantly more to right than left visual motion (t(16) = 3.74, p = 0.002 and t(16) = 2.91, p = .011 respectively, Fig. 6A). No subregions responded more to left than right visual motion. No subregions in the left hemisphere showed directional responses to visual motion (Fig. 6B). This motion selectivity was not seen in patients with vestibular neuritis (Fig. 6C-D). No subregions in either hemisphere showed activity that was affected by motion. The direct comparison of controls and patients showed significantly lower motion selectivity in vestibular neuritis patients than in controls for both R2 [lateral OP2] and R3 [posterior OP2] (R2: t(32) = 3.37, p = .002; R3: t(32) = 2.32, p = .027 respectively).

**Figure 6.**
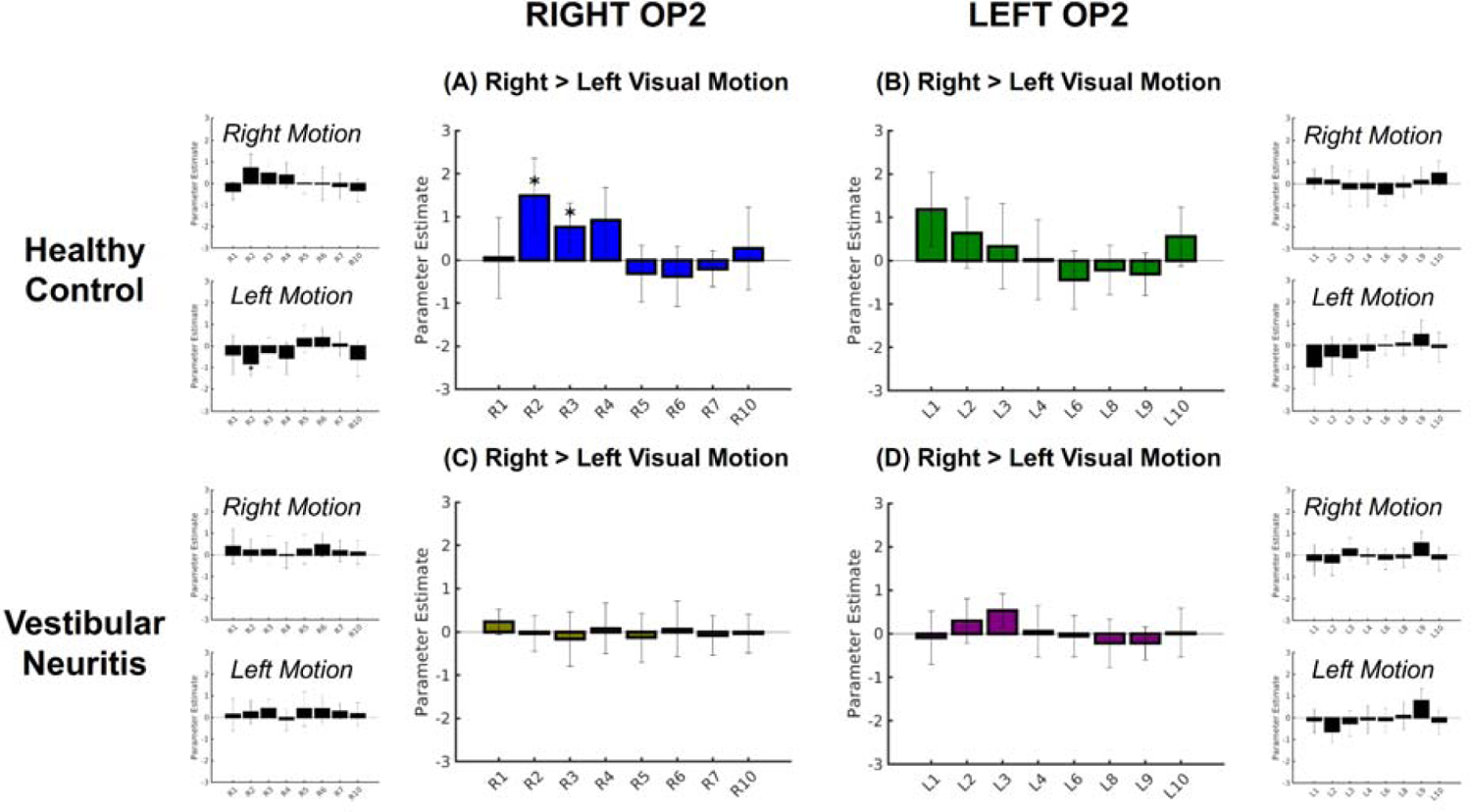
Right OP2 responds to visual motion direction in controls but not after vestibular neuritis. Larger bar charts show OP2 subregion responses for motion selectivity (right > left visual motion). Smaller bar charts show specific responses to right, and left motion (motion > static visual stimulation). Results for healthy controls (A, B) and patients with vestibular neuritis (C, D) are shown for right (A, C) and left (B, D) OP2. * indicates significant activation relative to baseline, FDR-corrected.

### Activation of right OP2 by caloric stimulation correlates with perceptions of dizziness, visual dependence and disability

Caloric irrigation of the left ear produced subjective feelings of dizziness in controls and vestibular neuritis patients. More dizziness was reported in patients than controls (control mean 3.11, S.D. 1.95; patient mean 4.01, S.D. 2.53; F(1,131) = 5.52, p = .02, Fig. 7A). Adjusted dizziness was determined, which is a fitted value for dizziness predicted by the caloric response with other predictors averaged out. In controls, adjusted dizziness correlated with activation in R3 [posterior OP2] (t(61) = 2.22, p = .030, Fig. 7A), but not other OP2 subregions which activated in either the right or left hemispheres. OP2 activation did not correlate with perceptions of dizziness in vestibular neuritis patients. Nystagmus during caloric stimulation can be quantified by its peak slow phase velocity (a measure of the vestibulo-ocular reflex). This did not differ between the groups (Fig. 7A) and did not correlate with dizziness in either group or with the activation of OP2 subregions during caloric stimulation.

**Figure 7.**
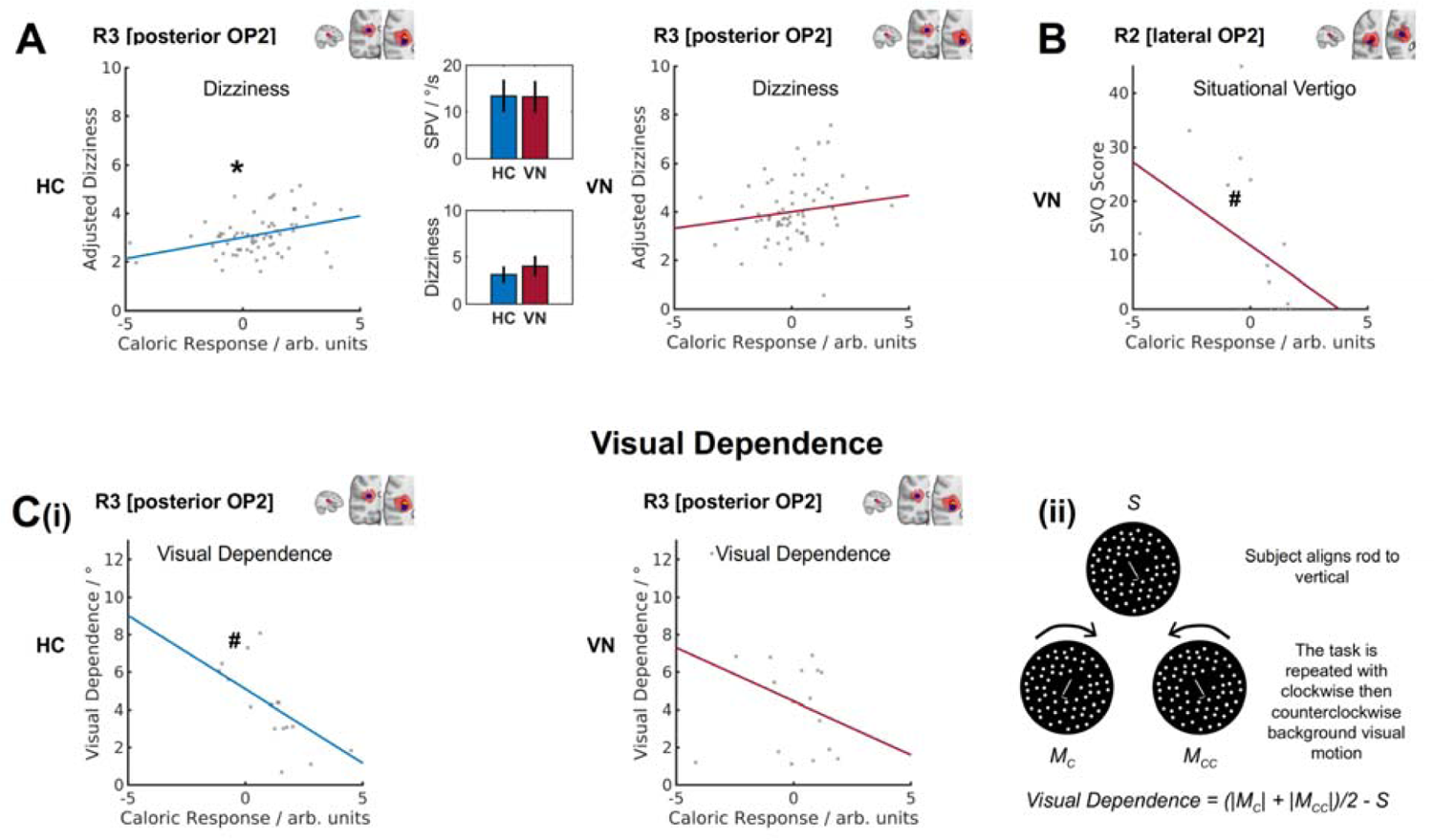
Correlation between subregion caloric response and dizziness, visual dependence and situational vertigo. (A) Scatter plots of dizziness and the caloric response in R3 [posterior OP2] in controls (HC) and patients with vestibular neuritis (VN); data points were obtained in each of four stimulus conditions. Inset bar charts show mean peak nystagmus slow phase velocity (SPV in degrees per second), and mean dizziness; error bars show the 95 % confidence intervals of the respective means. (B) Scatter plot of situational vertigo and R2 [lateral OP2] caloric activation in VN; the line of best fit is for illustration only. (C) (i) Scatter plots of visual dependence and R3 [posterior OP2] caloric response; (ii) illustration of rod and disc task from which visual dependence was calculated. The subjective visual vertical is measured when participants align a central rod overlying a static background (*S*); this is repeated during clockwise (*M_C_*) and counter-clockwise (*M_CC_*) rotation of the visual background; visual dependence is calculated as shown. * = p < .05 for caloric response predictor; # = p < .05 for Spearman correlation.

Individual differences in visual dependence also correlated with caloric responses in right posterior OP2. Visual dependence was similar in controls and patients with vestibular neuritis (control mean 3.90 °, S.D. 2.27 °, patient mean 4.57 °, S.D. 2.90 °). However, a negative correlation between R3 [posterior OP2] caloric activation and visual dependence was only significantly seen in controls (R3: Spearman r = −0.814, p < .001, Fig. 7C(i)). In patients with vestibular neuritis, OP2 activation did not significantly correlate with visual dependence (Fig. 7C(i)).

The functional impact of dizziness and vertigo in patients with vestibular neuritis was measured using the Dizziness Handicap Inventory, Situational Vertigo Questionnaire (SVQ) and Vertigo Symptom Scale. There was no significant relationship between these measures and caloric responses in R3 [posterior OP2], L3 [lateral OP2] or L10 [posterior OP2], which had activated in controls during caloric irrigation. We investigated whether a relationship existed in other subregions, which had similar whole brain connectivity: R1 [anterior OP2], R2 [lateral OP2] and L1 [anterior OP2] (see Fig. 4). Higher activity in R2 [lateral OP2] during caloric stimulation correlated significantly with lower SVQ scores (R2: Spearman r = −0.664, z = 2.99, p = .004, FDR-corrected p < .05, Fig. 7B). No other subregion responses correlated with SVQ scores. No significant correlations were found for DHI, or VSS scores and subregion caloric activity. Left OP2 caloric responses did not significantly correlate with any clinical symptom measures.

## Discussion

OP2 is a core region within the human cortical vestibular network on the basis of its location (Eickhoff et al., 2006b, 2006c), structural connectivity (Wirth et al., 2018; Indovina et al., 2020), and functional responses (Eickhoff et al., 2006c; zu Eulenburg et al., 2012). Here we define its functional anatomy by studying distinct patterns of connectivity measured using functional MRI.

The human cortical vestibular network is involved in higher order vestibular functions such as the perception of self-motion, judgements regarding verticality as well as balance control and spatial navigation (Lopez and Blanke, 2011). OP2 is well connected to this network (zu Eulenburg et al., 2012; Wirth et al., 2018; Indovina et al., 2020), and the area’s centrality in the network predicts the existence of subregions further connected to vestibular brain areas involved in processing higher-order visuo-vestibular functions.

Our results indeed showed that OP2 has strong functional connectivity to a range of regions involved in vestibular function including the primary somatosensory cortex, the parietal operculum, the supracalcarine cortex, the left inferior parietal lobule and the anterior cingulate cortex. For the first time, we identify a subregion within posterior OP2 (R3 [posterior OP2]) responsive to vestibular and visual information and with activity correlated with self-motion perception and visual dependence. The pattern of functional connectivity we observed is similar to that seen in the non-human primate PIVC (Guldin and Grüsser, 1998), a homologous area that also strongly connects to a number of cortical vestibular areas (Guldin et al., 1992; Guldin and Grüsser, 1998). Our human results align with those from non-human primates and suggest that OP2 and its analogue the PIVC act as cortical hubs for vestibular processing relevant to higher order vestibular functions (Guldin and Grüsser, 1998; Brandt and Dieterich, 1999; Lopez and Blanke, 2011).

The role of OP2 in processing visual motion information, however, has been contentious (Grüsser et al., 1990a; Brandt et al., 1998; Dieterich et al., 1998; Chen et al., 2010, 2016; Billington and Smith, 2015; Frank et al., 2016b). Key evidence against the processing of visual information stems from a study that showed no neural response in the macaque PIVC to moving dot stimuli (Chen et al., 2010). Chemical deactivation of anterior or posterior subregions of the macaque PIVC however impairs perceptual judgements of head orientation informed by visual cues (Chen et al., 2016), suggesting the PIVC processes visual motion information. In humans, parieto-insular areas including OP2 deactivate in response to visual motion (Kleinschmidt et al., 2002; Laurienti et al., 2002), particularly when it is the focus of attention (Frank et al., 2016a, 2020, 2021). Activation is instead seen in the retroinsular cortex (Brandt et al., 1998; Sunaert et al., 1999; Orban et al., 2003; Frank et al., 2014, 2016b). Here we showed a posterior subregion of right OP2 (R3 [posterior OP2]) extending into the anterior retroinsular cortex processes visual information, given its directional responses to visual motion stimuli and correlation between vestibular activity and visual dependence. Our results support the proposition that visuo-vestibular processing mainly occurs in the retroinsular region at the posterior border of OP2 (Frank and Greenlee, 2018), but also suggest this processing may extend into OP2.

There has also been debate about whether vestibular processing is lateralised in OP2 (Dieterich et al., 2003; Brandt and Dieterich, 2015; Kirsch et al., 2018; Wirth et al., 2018; Indovina et al., 2020; Raiser et al., 2020). The cytoarchitecture of OP2 is symmetrical (Eickhoff et al., 2006b). Studies of the region’s structural connectivity have shown mixed results with right (Wirth et al., 2018; Raiser et al., 2020), or left-lateralised patterns of cortico-cortical connectivity (Indovina et al., 2020). A functional connectivity study showed OP2 and neighbouring regions had both symmetric and lateralised cortico-cortical connectivity (Kirsch et al., 2018). Studies of cortical responses to vestibular stimuli have shown the right hemisphere (and thus right OP2) is dominant for vestibular functions (Dieterich et al., 2003; Dieterich and Brandt, 2008). We found no evidence of lateralisation in the pattern of functional connectivity in OP2 subregions (Fig. 4), or the response to caloric stimuli (Fig. 5). However, the effects of peripheral vestibular disease were asymmetrical and the relationship between activity and dizziness/visual dependence were only seen in right OP2 (Fig. 7A,C(i)), suggesting higher-order vestibular functions lateralise to the right hemisphere, as in handedness-related vestibular lateralization (Arshad et al., 2013, 2019; Chan et al., 2021).

Interestingly, OP2 responses to vestibular and visual stimuli were influenced by vestibular disease. In patients with vestibular neuritis, right OP2 responses to caloric stimulation (of the healthy ear) and visual stimuli were abolished, supporting a role for right OP2 in the normal processing of vestibular and visual stimuli. A recent study of cortical responses to galvanic stimuli in patients with bilateral vestibular failure found no relationship between dizziness handicap and activity in OP2 (Helmchen et al., 2019), and in our results the relationship between OP2 activity and visual dependence seen in healthy subjects was lost in vestibular neuritis patients. In addition, we observed that greater visually-induced dizziness (SVQ) in patients correlated with less vestibular activity in a distinct lateral subregion of right OP2 (R2 [lateral OP2]). This suggests that reduced vestibular responses in OP2 may be a marker of a poorer clinical outcome.

The change in OP2 activity in vestibular neuritis occurred despite similar vestibulo-ocular reflex function across patients (following stimulation of the healthy ear) and controls. These results align with previous work showing cortical vestibular processing can be modified separately from brainstem reflex function (Okada et al., 1999; Nigmatullina et al., 2015). Changes in cortical vestibular processing have been suggested to be relevant to adaptation and recovery following vestibular disease (Palla et al., 2008; Seemungal, 2014; Yip and Strupp, 2018). However, the mechanism underpinning reduced vestibular responses in OP2 to stimulation of the healthy ear is unclear. One possible mechanism is that there may be reduced vestibular input to OP2 following vestibular neuritis. This implies reduced vestibular signalling, separable from preserved vestibulo-ocular reflex functioning. Such reduced vestibular signalling may underpin the increase in visual dependence in patients. Animal studies have shown unilateral vestibular loss leads to distinct effects within the brainstem, and that the correlates of nystagmus and behavioural recovery differ (Smith and Curthoys, 1989). In contralesional medial vestibular nuclei, rapid restitution of resting neural activity occurs over a similar time frame to nystagmus recovery whereas desensitisation to vestibular signals recovers over a slower time frame in parallel with behavioral recovery (Markham et al., 1977; Yagi and Markham, 1984; Smith and Curthoys, 1988). To our knowledge, the cortical correlates of these changes have not previously been studied. Our findings in OP2 may be a correlate of such desensitisation.

Our study has a number of limitations. First, our comparisons operate on the assumption that OP2’s subregional anatomy is invariant between controls and patients. Reports of structural and functional changes in the vicinity of OP2 following peripheral vestibular disease mean this assumption may not have been satisfied (Helmchen et al., 2009, 2014). Group differences, if present, may have influenced correlations between neural activity in OP2 and vestibular functions such as self-motion perception and visual dependence. Second, OP2 activations in patients following caloric stimulation were weaker than in controls; this may have contributed to non-significant correlation between activation and higher vestibular function due to floor effects. Third, spatial smoothing in spatially constrained ICA may have increased the apparent spatial extent of functional subregions. Fourth, the results of our analyses depend on dimensionality in spatially constrained ICA. We maximised between-subject reproducibility to obtain an optimal number of dimensions for ICA in a data-driven way (Moher Alsady et al., 2016). In right OP2, the mean reproducibility curve had a global maximum at ten subregions. The data thus provided a clear justification for our choice. Notably, though a few subregions had similar whole-brain connectivity (Fig. 4), they nonetheless had distinct task-related activation, confirming good separability (Fig. 5, 6).

In summary, our results show a posterior subregion in right OP2 (R3 [posterior OP2]) is well connected to other areas in the cortical vestibular network, and the subregion processes visual and vestibular information relevant to the perception of self-motion and verticality. We also find that vestibular responses in a different subregion (R2 [lateral OP2]) are variably affected by chronic vestibular neuritis, and this effect appears relevant to symptomatic recovery.

## Acknowledgements

This work was supported by funding from the UK Medical Research Council (MR/J004685/1), the Dunhill Medical Trust (R481/0516) and the Imperial National Institute for Health Research (NIHR) Biomedical Research Centre. D.K is supported by the NIHR University College London Hospitals Biomedical Research Centre. D.J.S is supported by the UK Dementia Research Institute Care Research & Technology Centre, and the Centre for Injury studies at Imperial College London. The authors would like to thank Dr. Edward Richard Roberts and colleagues who collected the data re-analysed in this study (Roberts et al., 2018). We also thank Dr. Christophe Lopez, Aix-Marseille Université (Marseille, France) for interim data from a meta-analysis of brain responses to vestibular stimuli (Lopez et al., 2012).

## Abbreviated title

Human vestibular cortex functional anatomy

## References

1. Arshad Q, Nigmatullina Y, Bronstein AM (2013) Handedness-related cortical modulation of the vestibular-ocular reflex. J Neurosci 33:3221–3227.

2. Arshad Q, Ortega MC, Goga U, Lobo R, Siddiqui S, Mediratta S, Bednarczuk NF, Kaski D, Bronstein AM (2019) Interhemispheric control of sensory cue integration and self-motion perception. Neuroscience 408:378–387.

3. Beckmann CF, Jenkinson M, Smith SM (2003) General multilevel linear modeling for group analysis in FMRI. Neuroimage 20:1052–1063.

4. Billington J, Smith AT (2015) Neural mechanisms for discounting head-roll-induced retinal motion. J Neurosci 35:4851–4856.

5. Braga RM, Sharp DJ, Leeson C, Wise RJS, Leech R (2013) Echoes of the brain within default mode, association, and heteromodal cortices. J Neurosci 33:14031–14039.

6. Brandt T, Bartenstein P, Janek A, Dieterich M (1998) Reciprocal inhibitory visual-vestibular interaction. Visual motion stimulation deactivates the parieto-insular vestibular cortex. Brain 121:1749–1758.

7. Brandt T, Dieterich M (1999) The vestibular cortex. Its locations, functions, and disorders. Ann N Y Acad Sci 871:293–312.

8. Brandt T, Dieterich M (2015) Does the vestibular system determine the lateralization of brain functions? J Neurol 262:214–215.

9. Bronstein AM, Dieterich M (2019) Long-term clinical outcome in vestibular neuritis. Curr Opin Neurol 32:174–180.

10. Chan YM, Wong Y, Khalid N, Wastling S, Flores-Martin A, Frank L-A, Koohi N N, Arshad Q, Davagnanam I, Kaski D (2021) Prevalence of acute dizziness and vertigo in cortical stroke. Eur J Neurol Available at: http://dx.doi.org/10.1111/ene.14964.

11. Chen A, DeAngelis GC, Angelaki DE (2010) Macaque parieto-insular vestibular cortex: responses to self-motion and optic flow. J Neurosci 30:3022–3042.

12. Chen A, Gu Y, Liu S, DeAngelis GC, Angelaki DE (2016) Evidence for a Causal Contribution of Macaque Vestibular, But Not Intraparietal, Cortex to Heading Perception. J Neurosci 36:3789–3798.

13. Cousins S, Cutfield NJ, Kaski D, Palla A, Seemungal BM, Golding JF, Staab JP, Bronstein AM (2014) Visual dependency and dizziness after vestibular neuritis. PLoS One 9:e105426.

14. Cullen KE (2019) Vestibular processing during natural self-motion: implications for perception and action. Nat Rev Neurosci 20:346–363.

15. De Simoni S, Jenkins PO, Bourke NJ, Fleminger JJ, Hellyer PJ, Jolly AE, Patel MC, Cole JH, Leech R, Sharp DJ (2018) Altered caudate connectivity is associated with executive dysfunction after traumatic brain injury. Brain 141:148–164.

16. Dieterich M, Bense S, Lutz S, Drzezga A, Stephan T, Bartenstein P, Brandt T (2003) Dominance for vestibular cortical function in the non-dominant hemisphere. Cereb Cortex 13:994–1007.

17. Dieterich M, Brandt T (2008) Functional brain imaging of peripheral and central vestibular disorders. Brain 131:2538–2552.

18. Dieterich M, Bucher SF, Seelos KC, Brandt T (1998) Horizontal or vertical optokinetic stimulation activates visual motion-sensitive, ocular motor and vestibular cortex areas with right hemispheric dominance. An fMRI study. Brain 121 ( Pt 8):1479–1495.

19. Eickhoff SB, Heim S, Zilles K, Amunts K (2006a) Testing anatomically specified hypotheses in functional imaging using cytoarchitectonic maps. Neuroimage 32:570–582.

20. Eickhoff SB, Schleicher A, Zilles K, Amunts K (2006b) The human parietal operculum. I. Cytoarchitectonic mapping of subdivisions. Cereb Cortex 16:254–267.

21. Eickhoff SB, Weiss PH, Amunts K, Fink GR, Zilles K (2006c) Identifying human parieto-insular vestibular cortex using fMRI and cytoarchitectonic mapping. Hum Brain Mapp 27:611–621.

22. Frank SM, Baumann O, Mattingley JB, Greenlee MW (2014) Vestibular and visual responses in human posterior insular cortex. J Neurophysiol 112:2481–2491.

23. Frank SM, Forster L, Pawellek M, Malloni WM, Ahn S, Tse PU, Greenlee MW (2021) Visual Attention Modulates Glutamate-Glutamine Levels in Vestibular Cortex: Evidence from Magnetic Resonance Spectroscopy. J Neurosci 41:1970–1981.

24. Frank SM, Greenlee MW (2018) The parieto-insular vestibular cortex in humans: more than a single area? J Neurophysiol 120:1438–1450.

25. Frank SM, Pawellek M, Forster L, Langguth B, Schecklmann M, Greenlee MW (2020) Attention Networks in the Parietooccipital Cortex Modulate Activity of the Human Vestibular Cortex during Attentive Visual Processing. J Neurosci 40:1110–1119.

26. Frank SM, Sun L, Forster L, Peter UT, Greenlee MW (2016a) Cross-modal attention effects in the vestibular cortex during attentive tracking of moving objects. Journal of Neuroscience 36:12720–12728.

27. Frank SM, Wirth AM, Greenlee MW (2016b) Visual-vestibular processing in the human Sylvian fissure. J Neurophysiol 116:263–271.

28. Genovese CR, Lazar NA, Nichols T (2002) Thresholding of statistical maps in functional neuroimaging using the false discovery rate. Neuroimage 15:870–878.

29. Glasser MF, Sotiropoulos SN, Wilson JA, Coalson TS, Fischl B, Andersson JL, Xu J, Jbabdi S, Webster M, Polimeni JR, Van Essen DC, Jenkinson M, WU-Minn HCP Consortium (2013) The minimal preprocessing pipelines for the Human Connectome Project. Neuroimage 80:105–124.

30. Grüsser OJ, Pause M, Schreiter U (1990a) Vestibular neurones in the parieto-insular cortex of monkeys (Macaca fascicularis): visual and neck receptor responses. J Physiol 430:559–583.

31. Grüsser OJ, Pause M, Schreiter U (1990b) Localization and responses of neurones in the parieto-insular vestibular cortex of awake monkeys (Macaca fascicularis). J Physiol 430:537–557.

32. Guldin WO, Akbarian S, Grüsser OJ (1992) Cortico-cortical connections and cytoarchitectonics of the primate vestibular cortex: a study in squirrel monkeys (Saimiri sciureus). J Comp Neurol 326:375–401.

33. Guldin WO, Grüsser OJ (1998) Is there a vestibular cortex? Trends Neurosci 21:254–259.

34. Halmagyi GM, Curthoys IS (1988) A clinical sign of canal paresis. Arch Neurol 45:737–739.

35. Hastie T, Tibshirani R, Friedman J (2009) Hierarchical clustering. The elements of statistical learning 2.

36. Helmchen C, Klinkenstein J, Machner B, Rambold H, Mohr C, Sander T (2009) Structural changes in the human brain following vestibular neuritis indicate central vestibular compensation. Ann N Y Acad Sci 1164:104–115.

37. Helmchen C, Rother M, Spliethoff P, Sprenger A (2019) Increased brain responsivity to galvanic vestibular stimulation in bilateral vestibular failure. Neuroimage Clin 24:101942.

38. Helmchen C, Ye Z, Sprenger A, Münte TF (2014) Changes in resting-state fMRI in vestibular neuritis. Brain Struct Funct 219:1889–1900.

39. Himberg J, Hyvärinen A, Esposito F (2004) Validating the independent components of neuroimaging time series via clustering and visualization. Neuroimage 22:1214–1222.

40. Imate Y, Sekitani T (1993) Vestibular compensation in vestibular neuronitis. Long-term follow-up evaluation. Acta Otolaryngol 113:463–465.

41. Indovina I, Bosco G, Riccelli R, Maffei V, Lacquaniti F, Passamonti L, Toschi N (2020) Structural connectome and connectivity lateralization of the multimodal vestibular cortical network. Neuroimage 222:117247.

42. Jacob RG, Lilienfeld SO, Furman JMR, Durrant JD, Turner SM (1989) Panic disorder with vestibular dysfunction: Further clinical observations and description of space and motion phobic stimuli. J Anxiety Disord 3:117–130.

43. Jacobson GP, Newman CW (1990) The development of the Dizziness Handicap Inventory. Arch Otolaryngol Head Neck Surg 116:424–427.

44. Jenkinson M, Bannister P, Brady M, Smith S (2002) Improved optimization for the robust and accurate linear registration and motion correction of brain images. Neuroimage 17:825–841.

45. Kanayama R, Bronstein AM, Gresty MA, Brookes GB, Faldon ME, Nakamura T (1995) Perceptual studies in patients with vestibular neurectomy. Acta Otolaryngol Suppl 520 Pt 2:408–411.

46. Kirsch V, Boegle R, Keeser D, Kierig E, Ertl-Wagner B, Brandt T, Dieterich M (2018) Handedness-dependent functional organizational patterns within the bilateral vestibular cortical network revealed by fMRI connectivity based parcellation. Neuroimage 178:224–237.

47. Kleinschmidt A, Thilo KV, Büchel C, Gresty MA, Bronstein AM, Frackowiak RSJ (2002) Neural correlates of visual-motion perception as object- or self-motion. Neuroimage 16:873–882.

48. Laurienti PJ, Burdette JH, Wallace MT, Yen Y-F, Field AS, Stein BE (2002) Deactivation of sensory-specific cortex by cross-modal stimuli. J Cogn Neurosci 14:420–429.

49. Leech R, Braga R, Sharp DJ (2012) Echoes of the brain within the posterior cingulate cortex. J Neurosci 32:215–222.

50. Leech R, Kamourieh S, Beckmann CF, Sharp DJ (2011) Fractionating the default mode network: distinct contributions of the ventral and dorsal posterior cingulate cortex to cognitive control. J Neurosci 31:3217–3224.

51. Lopez C, Blanke O (2011) The thalamocortical vestibular system in animals and humans. Brain Res Rev 67:119–146.

52. Lopez C, Blanke O, Mast FW (2012) The human vestibular cortex revealed by coordinate-based activation likelihood estimation meta-analysis. Neuroscience 212:159–179.

53. Markham CH, Yagi T, Curthoys IS (1977) The contribution of the contralateral labyrinth to second order vestibular neuronal activity in the cat. Brain Res 138:99–109.

54. MATLAB (2021) Adjusted response plot of linear regression model - MATLAB plotAdjustedResponse. Available at: https://www.mathworks.com/help/stats/linearmodel.plotadjustedresponse.html [Accessed May 15, 2021].

55. Moher Alsady T, Blessing EM, Beissner F (2016) MICA-A toolbox for masked independent component analysis of fMRI data. Hum Brain Mapp 37:3544–3556.

56. Nickerson LD, Smith SM, Öngür D, Beckmann CF (2017) Using Dual Regression to Investigate Network Shape and Amplitude in Functional Connectivity Analyses. Front Neurosci 11:115.

57. Nigmatullina Y, Hellyer PJ, Nachev P, Sharp DJ, Seemungal BM (2015) The neuroanatomical correlates of training-related perceptuo-reflex uncoupling in dancers. Cereb Cortex 25:554–562.

58. Okada T, Grunfeld E, Shallo-Hoffmann J, Bronstein AM (1999) Vestibular perception of angular velocity in normal subjects and in patients with congenital nystagmus. Brain 122 ( Pt 7):1293–1303.

59. Orban GA, Fize D, Peuskens H, Denys K, Nelissen K, Sunaert S, Todd J, Vanduffel W (2003) Similarities and differences in motion processing between the human and macaque brain: evidence from fMRI. Neuropsychologia 41:1757–1768.

60. Palla A, Straumann D, Bronstein AM (2008) Vestibular neuritis: vertigo and the high-acceleration vestibulo-ocular reflex. J Neurol 255:1479–1482.

61. Pruim RHR, Mennes M, van Rooij D, Llera A, Buitelaar JK, Beckmann CF (2015) ICA-AROMA: A robust ICA-based strategy for removing motion artifacts from fMRI data. Neuroimage 112:267–277.

62. Raiser TM, Flanagin VL, Duering M, van Ombergen A, Ruehl RM, Zu Eulenburg P (2020) The human corticocortical vestibular network. Neuroimage 223:117362.

63. Roberts RE, Ahmad H, Arshad Q, Patel M, Dima D, Leech R, Seemungal BM, Sharp DJ, Bronstein AM (2017) Functional neuroimaging of visuo-vestibular interaction. Brain Struct Funct 222:2329–2343.

64. Roberts RE, Ahmad H, Patel M, Dima D, Ibitoye R, Sharif M, Leech R, Arshad Q, Bronstein AM (2018) An fMRI study of visuo-vestibular interactions following vestibular neuritis. Neuroimage Clin 20:1010–1017.

65. Salimi-Khorshidi G, Douaud G, Beckmann CF, Glasser MF, Griffanti L, Smith SM (2014) Automatic denoising of functional MRI data: Combining independent component analysis and hierarchical fusion of classifiers. NeuroImage 90:449–468 Available at: http://dx.doi.org/10.1016/j.neuroimage.2013.11.046.

66. Seemungal BM (2014) The cognitive neurology of the vestibular system. Curr Opin Neurol 27:125–132.

67. Smith PF, Curthoys IS (1988) Neuronal activity in the ipsilateral medial vestibular nucleus of the guinea pig following unilateral labyrinthectomy. Brain Res 444:308–319.

68. Smith PF, Curthoys IS (1989) Mechanisms of recovery following unilateral labyrinthectomy: a review. Brain Res Brain Res Rev 14:155–180.

69. Smith SM (2002) Fast robust automated brain extraction. Hum Brain Mapp 17:143–155.

70. Smith SM, Fox PT, Miller KL, Glahn DC, Fox PM, Mackay CE, Filippini N, Watkins KE, Toro R, Laird AR, Beckmann CF (2009) Correspondence of the brain’s functional architecture during activation and rest. Proc Natl Acad Sci U S A 106:13040–13045.

71. Sunaert S, Van Hecke P, Marchal G, Orban GA (1999) Motion-responsive regions of the human brain. Exp Brain Res 127:355–370.

72. Wirth AM, Frank SM, Greenlee MW, Beer AL (2018) White Matter Connectivity of the Visual-Vestibular Cortex Examined by Diffusion-Weighted Imaging. Brain Connect 8:235–244.

73. Woolrich MW, Ripley BD, Brady M, Smith SM (2001) Temporal autocorrelation in univariate linear modeling of FMRI data. Neuroimage 14:1370–1386.

74. Yagi T, Markham CH (1984) Neural correlates of compensation after hemilabyrinthectomy. Exp Neurol 84:98–108.

75. Yang Z, LaConte S, Weng X, Hu X (2008) Ranking and averaging independent component analysis by reproducibility (RAICAR). Hum Brain Mapp 29:711–725.

76. Yardley L, Masson E, Verschuur C, Haacke N, Luxon L (1992) Symptoms, anxiety and handicap in dizzy patients: development of the vertigo symptom scale. J Psychosom Res 36:731–741.

77. Yip CW, Strupp M (2018) The Dizziness Handicap Inventory does not correlate with vestibular function tests: a prospective study. J Neurol 265:1210–1218.

78. zu Eulenburg P, Caspers S, Roski C, Eickhoff SB (2012) Meta-analytical definition and functional connectivity of the human vestibular cortex. Neuroimage 60:162–169.

